# Assessment of Knowledge, Attitudes, Practices, and Perceptions Regarding Use of Sunscreen Among Pharmacists in Jordan

**DOI:** 10.1101/2025.10.23.25338664

**Authors:** Alaa Alghananim, Roua Aldala’een, Abdallah Y Naser, Alaa Ahmed Alsharif, Sarah Khamis

**Affiliations:** Department of pharmaceutical sciences, Faculty of Pharmacy, Jerash university, Jerash, Jordan; Department of Applied Pharmaceutical Sciences and Clinical Pharmacy, Faculty of Pharmacy, Isra University, Amman, Jordan; Department of Pharmacy Practice, College of pharmacy, Princess Nourah bint Abdulrahman University

## Abstract

Sunscreens are the agents to protect the skin from the destructive effects of the Ultraviolet radiation. Pharmacists are the expert healthcare providers in guiding the consumers regarding usage of sunscreen agents. this study is to evaluate the extent to which pharmacists in Jordan are aware of the knowledge, attitudes, practices, and perceptions of sunscreen usage.

A cross-sectional study was conducted among the pharmacists in Jordan using a self-administered questionnaire which was created using Google Forms^®^ and was distributed online on different social media platforms. The questionnaire containing 5 sections to assess demographics (9 items), knowledge (11 items), attitude (8 items), practice (12 items), and perception (2 items). The data was subjected to descriptive and inference statistics.

A total of 407 pharmacists working in different fields completed the survey, consisted of (79.6%) female and 20.4% male. 56.5% of participants were less than 30 years old, while (56%) of participants have experience of less than 5 years. The results showed that knowledge, attitude and practice scores were strongly impacted by age (p=0.011), (p<0.001) respectively. In addition, (53.8%) were familiar with Fitzpatrick scale of skin type that categorizes the skin into 6 types, and 232 (57.0%) responded correctly regarding the active ingredient’s classification of sunscreen agents. Also, knowledge score (p=0.023), attitude and practice score (p=0.007) were significantly influenced by years of experience. Furthermore, (55.3%) and (66.6%) of pharmacists correctly identified that gel form and cream form are the recommended forms for clients of oily skin and dry skin respectively.

In conclusion, the knowledge and practice of sunscreen use among pharmacists in Jordan were sub optimal. This demonstrated that pharmacist needs more specialized courses during practice for their continuing professional development to enhance the knowledge and the practice.

## Introduction

Sunscreens are considered photoprotection agents which are formulated to protect the skin from the harmful effects of solar radiation, particularly the UV radiation, either by its ability to absorb, or scatter solar rays. [1]

UV radiation is subdivided into UVA, UVB and UVC. UVA radiation (315–400 nm) produces an excess of reactive oxygen species, which in turn indirectly damage DNA. While UVB radiation (280-320nm) can directly damage DNA by forming pyrimidine dimers, which can trigger apoptosis or lead to DNA replication errors, resulting in mutations and potentially cancer. On the other hand, UVC radiation is blocked by the ozone layer in the atmosphere, preventing it from reaching the Earth’s surface. [2]

Sunscreen agents which absorb UV radiation and convert it into an insignificant amount of heat are classified as organic or chemical sunscreens while sunscreen agents which reflect, and scatter light are classified as inorganic or physical sunscreens. [3]

The effectiveness of sunscreen in providing photoprotection is determined by sun protection factor (SPF) which measure a sunscreen ability to protect one from sunburn primarily resulting from UVB and also by the protection grade of UVA (PA) values. [4] The Sun Protection Factor (SPF), as defined by the FDA in 1978, as the numerical ratio between the minimal erythemal dose (MED) of sunscreen-protected skin, applied at a rate of 2 mg/cm², and the minimal erythemal dose of unprotected skin. [5]

Since the 1980s, international sun protection campaigns have played a crucial role in raising awareness about the risks of excessive sun exposure and the importance of sunscreens. [6] It was found that sunscreen agents play a crucial role in decreasing the prevalence of skin disorders in humans caused by UV rays such as sunburn, skin aging, pigment symptoms and also play a role in the prevention of epithelial and nonepithelial skin cancer. [7]

Many sunscreens topical formulation types are available in pharmaceutical market including gel, cream, lotion, foam and stick.

Sunscreen agents are classified as cosmetics product according to EU regulations while they are over the counter (OTC) drugs according to US regulations [8], consequently pharmacists are key healthcare providers who play an essential role in medication and cosmetics management. [9] Pharmacists can guide individuals in making informed decisions regarding usage of sunscreen agents.

The objective of this study is to evaluate the extent to which pharmacists in Jordan are aware of the knowledge, attitudes, practices, and perceptions of sunscreen usage.

## Method

### Study design and setting

This is a cross-sectional questionnaire-based study among pharmacists in Jordan conducted between 21^st^ January to 8^th^ February 2025. The study was conducted through an online questionnaire, developed using Google Forms^®^. The researchers shared the form link on social media platforms to distribute it to ensure a wide reach.

### Ethics and consent

All individuals were informed that the study was voluntary through the description provided on the Google form® questionnaire, and consent was attained from all participants. The questionnaire was developed based on insights from earlier studies on sunscreen agents use, the study received approval from the Deanship of Scientific Research and Graduate Studies at Jerash University, with the ethical approval number (2025/2024/2/7). To ensure anonymity, no personally identifiable information was collected during the data collection process.

### Questionnaire

The design of the questionnaire included sections on knowledge of sunscreen agents, practices related to sunscreen use, attitudes, and perceptions. The questionnaire was organized into five sections:

- Demographic Information: This section included questions about the participants’ age, gender, degree, years of experience, current workplace, graduate university, and grade point average.
- Knowledge: Comprised of eleven questions focused on assessing pharmacists’ knowledge of sunscreen agents, including basic information about their effectiveness, ingredients, and proper use.
- Attitude: Contained eight questions aimed at understanding pharmacists’ beliefs towards sunscreen use
- Practice: This section consisted of twelve questions that primarily examined the actual behaviors of pharmacists regarding sunscreen use and their recommendations for patients
- Perception: Included two questions designed to assess pharmacists’ opinions and understanding of sunscreen.

Content validity was conducted among three experienced academics, to assure that all the questions are reliable and pertinent to the pharmacists.

According to statistics by the Jordanian Pharmaceutical Association (JPA) report a total of 34325 registered pharmacists in 2024 [10], Thus, the minimum required sample size which was determined using SurveyMonkey method was 380 for (95%) confidence interval and (5%) margin of error.

A total of 407 complete responses were obtained and considered in this study.

### Statistical analysis

The data was subjected to descriptive and inference statistics. Results are revealed as a Mean ± SD. For analyzing data used Pearson’s chi-square, Pearson correlation coefficient, and independent t-tests by Statistical Package for Social Sciences (SPSS) software version 26 for Windows. The level of statistical significance was set at p < 0.05 (two-sided p-values).

## Results

### Demographic date

The total number of pharmacists who completed the survey is 407 pharmacists working in different fields, the study sample consisted of (79.6%) female and (20.4%) male. (56.5%) of participants were less than 30 years old and a total of (77.1%) of the participants held a bachelor’s degree in pharmacy. (56%) of participants have experience of less than 5 years and (59.7%) of participants working in middle Jordan region. Table 1 describes the detailed demographic date of pharmacist’s participants in the survey.

**Table 1.** Demographic Date of Participants.

| Factor | N (%) |
| --- | --- |
| **Age:** |  |
| Less than 30 years | 230 (56.5%) |
| 30-39 years | 126 (31.0%) |
| 40-49 years | 38 (9.3%) |
| 50-59 years | 12 (2.9%) |
| Above 60 years | 1 (0.2%) |
| **Gender:** |  |
| Male | 324 (79.6%) |
| Female | 83 (20.4%) |
| **Degree:** |  |
| BSc. in Pharmacy | 314 (77.1%) |
| BSc. in Doctor of Pharmacy (PharmD) | 43 (10.6%) |
| Postgraduate (MSc. / Ph.D.) | 50 (12.3%) |
| **Years of Experience:** |  |
| Less than 5 years | 228 (56.0%) |
| 5-10 years | 105 (25.8%) |
| 11-15 years | 42 (10.3%) |
| More than 15 years | 32 (7.9%) |
| **Current Workplace:** |  |
| Chain pharmacy | 71 (17.4%) |
| Independent community pharmacy | 114 (28.0%) |
| Hospital | 47 (11.5%) |
| Medical representative | 15 (3.7%) |
| Pharmacist at Ministry of Health | 28 (6.9 %) |
| Academia | 41 (10.1%) |
| others | 91 (22.4%) |
| **Working Area:** |  |
| North Jordan (Irbid, Jerash, Ajlun, Mafraq) | 130 (31.9%) |
| Middle Jordan (Amman, Balqa, Madaba, Zarqa) | 243 (59.7%) |
| South Jordan (Aqaba, Karak. Ma'an, Tafilah) | 34 (8.4%) |
| **Status:** |  |
| Single | 222 (54.5%) |
| Married | 180 (44.2%) |
| Other | 5 (1.2%) |
| **Graduate University:** |  |
| Public university | 182 (44.7%) |
| Private university | 225 (55.3%) |

### Knowledge of Sun Protection Factor (SPF)

Knowledge score (p=0.011) was strongly impacted by age, with the 60+ age group having the highest knowledge. Also, knowledge (p=0.023) was significantly influenced by years of experience; pharmacists with less experience demonstrated superior knowledge. In addition, knowledge was influenced by current employment (p=0.017), with Ministry of Health pharmacists ranking the lowest and independent community pharmacists the highest. Furthermore, knowledge (p=0.043) was influenced by marital status, with singles having higher knowledge.

Out of 407 pharmacists, (70.5%) were familiar with the concept of Sun Protection Factor (SPF) and (47.7%) responded that SPF is a measure of the level of protection against UVB rays. (75.9%) answered correctly that SPF value determines how long it would take for your skin to burn with sunscreen compared to without the use. (30.2%) correctly identified that SPF 30 level provides (97%) protection.

Table 2 summarizes the responses of pharmacists regarding questions related to knowledge of Sun Protection Factor (SPF)

**Table 2.** Questionnaire Results Regarding Knowledge of Sun Protection Factor (SPF).

| Question | N (%) |
| --- | --- |
| **How familiar are you with the term SPF (sun protection factor)?** |  |
| Very familiar | 1 (0.2%) |
| Somewhat familiar | 103 (25.3%) |
| Familiar | 268 (70.3%) |
| Not familiar at all | 17 (4.2) |
| **Which of the following statements about SPF is correct?** |  |
| SPF measure the level of protection against UVA rays | 181 (44.5%) |
| SPF measure the level of protection against UVB rays | 194 (47.7%) |
| SPF measure the level of protection against UVC rays | 12 (2.9%) |
| I don't know | 20 (4.9%) |
| **SPF determine how long it would take for your skin to burn with sunscreen compared to without the use** |  |
| Yes | 309 (75.9%) |
| No | 98 (24.1%) |
| **Which SPF level provides 97% protection?** |  |
| SPF 15 | 36 (8.8%) |
| SPF 30 | 123 (30.2%) |
| SPF 50 | 220 (54.1%) |
| I don't know | 28 (6.9%) |
| **For patients who are concerned about skin irritation, do you generally recommend: (Select all that apply)** |  |
| Mineral sunscreens (zinc oxide, titanium dioxide) | 217 (53.3%) |
| Fragrance-free, Alcohol-free sunscreens | 250 (61.4%) |
| Sunscreens with added skincare benefits (e.g., moisturizing agents) | 202 (49.6%) |
| Sunscreen with SPF 90 or more | 68 (16.7%) |
| I'm not sure | 47 (11.5%) |

### Knowledge of sunscreen types and Fitzpatrick skin types

The results showed that pharmacists in Jordan recognize the difference between mineral (physical) sunscreen and chemical sunscreen, (74.9%) responded that the sunscreen agent which works by scatter or reflects UV radiation is the mineral (physical) sunscreen and (70.0%) responded that the sunscreen agent which works by changing UV radiation into heat, then release heat from skin is the chemical sunscreen. When the pharmacists were asked about the active ingredients for each physical and chemical sunscreen (multiple responses were allowed), 81 (20%) pharmacist answered complete correct answer regarding active ingredients which are classified as physical sunscreen which were zinc oxide, titanium oxide and talc. While 151 (37.0%) answered the correct answer for active ingredients classified as chemical sunscreen which were octocrylene, ethylhexyl methoxycinnamate and avobenzone.

Out of 407 pharmacists, 219 (53.8%) were familiar with Fitzpatrick scale of skin type that categorizes the skin into 6 types. In this study (33.9%) responded correctly to the number of skin types.

Participants’ knowledge of sunscreen types and Fitzpatrick scale was influenced by age, years of experience, current workplace, and marital status (p<0.05). Participants aged less than 30 years showed higher knowledge score compared to other. Those who have an experience of less than 5 years, single and working in independent community pharmacy showed similar findings.

Table 3 summarizes the responses of pharmacists regarding questions related to knowledge of sunscreen active ingredients and Fitzpatrick scale.

**Table 3.** Questionnaire Results Regarding Sunscreen Knowledge of sunscreen types and Fitzpatrick scale.

| Question | N (%) |
| --- | --- |
| **The sunscreen agent which works by scatter or reflects UV radiation is;** |  |
| Mineral (Physical) sunscreen | 305 (74.9%) |
| Chemical sunscreen | 77 (18.9%) |
| I don't know | 25 (6.1%) |
| **The sunscreen agent which works by changing UV radiation into heat, then release heat from skin is;** |  |
| Mineral (Physical) sunscreen | 285 (70.0%) |
| Chemical sunscreen | 95 (23.3%) |
| I don't know | 27 (6.6%) |
| **Which of the following ingredients are classified as physical sunscreens?**<br>**(you may choose more than one answer)** |  |
| Zinc oxide | 274 (67.3%) |
| Titanium dioxide | 245 (60.2%) |
| Talc | 78 (19.2%) |
| Octocrylene | 29 (7.1%) |
| Ethylhexyl methoxycinnamate | 34 (8.4%) |
| Avobenzone | 31 (7.6%) |
| I don't know | 60 (14.7%) |
| **Which of the following ingredients are classified as chemical sunscreens? (you may choose more than one answer)** |  |
| Zinc oxide | 70 (17.2%) |
| Titanium dioxide | 66 (16.2%) |
| Talc | 50 (12.3%) |
| Octocrylene | 180 (44.3%) |
| Ethylhexyl methoxycinnamate | 171 (42%) |
| Avobenzone | 176 (43.2%) |
| I don't know | 76 (18.7%) |
| **Are you familiar with Fitzpatrick skin types of classification?** |  |
| Yes | 219 (53.8%) |
| No | 188 (46.2%) |
| **How many skin types are categorized according to the Fitzpatrick scale?** |  |
| 2 | 19 (4.7%) |
| 4 | 89 (21.9%) |
| 6 | 138 (33.9%) |
| 10 | 13 (3.2%) |
| I don't know | 148 (36.4%) |
| **Do you know your skin type according to the Fitzpatrick scale?** |  |
| Yes | 185 (45.5%) |
| No | 90 (22.1%) |
| I'm not sure | 132 (32.4%) |

### Attitude and practice of sunscreen usage

Table 4 summarizes the responses of pharmacists regarding attitude and practice of sunscreen usage.

**Table 4.** Questionnaire Results Regarding Attitude and Practice of sunscreens usage.

| Question | N (%) |
| --- | --- |
| **Do you think that sunscreens are effective in sunburn prevention?** |  |
| Yes | 385 (94.6%) |
| No | 22 (5.4%) |
| **Do you think that sunscreens can lead to vitamin D deficiency?** |  |
| Yes | 177 (43.5%) |
| No | 230 (56.5%) |
| **Do you believe that sunscreens cause skin irritation?** |  |
| Yes | 233 (57.2%) |
| No | 174 (42.8%) |
| **Do you believe that sunscreens are not effective in preventing skin damage?** |  |
| Yes | 113 (27.8%) |
| No | 294 (72.2%) |
| **Are sunscreens safe to use during pregnancy?** |  |
| Yes | 365 (89.7%) |
| No | 42 (10.3%) |
| **Are sunscreens safe for babies older than 6 months old?** |  |
| Yes | 286 (70.3%) |
| No | 121 (29.7%) |
| **Do you believe that sunscreens only need to be applied once daily?** |  |
| Yes | 322 (79.1%) |
| No | 85 (20.9%) |
| **Do you apply sunscreen for yourself?** |  |
| Yes | 350 (86.0%) |
| No | 57 (14.0%) |
| **Do you recommend using sunscreen to others? (relatives, friends or patients)** |  |
| Yes | 384 (94.3%) |
| No | 23 (5.7%) |
| **When do you recommend users apply chemical sunscreen?** |  |
| 15-30 minutes before sun exposure | 332 (81.6%) |
| Immediately before going outside | 52 (12.8%) |
| I don't pay attention to timing | 23 (5.7%) |
| **How often do you recommend users apply sunscreen throughout the day?** |  |
| Once a day | 73 (17.9%) |
| Every 2-4 hours | 294 (72.2%) |
| Only when they are outdoors in a sunny day | 40 (9.8%) |
| **What SPF do you usually recommend?** |  |
| SPF 15 | 35 (8.6%) |
| SPF 30 | 115 (28.3%) |
| SPF 50 | 257 (63.1%) |
| **What type of sunscreen do you recommend mostly?** |  |
| Mineral (Physical) sunscreen | 196 (48.2%) |
| Chemical sunscreen | 110 (27.0%) |
| No preferences | 101 (24.8%) |
| **How often do you explain the differences between sunscreen formulations?**<br>**(e.g., mineral vs. chemical, lotion vs. spray)** |  |
| Always | 114 (28.0%) |
| Often | 144 (35.4%) |
| Occasionally | 63 (15.5%) |
| Rarely | 51 (12.5%) |
| Never | 35 (8.6%) |
| **What factors influence your recommendation of a sunscreen**<br>**formulation? (Select all that apply)** |  |
| Patient's skin type (e.g., oily, dry, sensitive) | 364 (85.0%) |
| Patient's lifestyle or activity level (e.g., sports, beach, everyday use) | 247 (60.7%) |
| Ease of application | 202 (49.6%) |
| Skin concerns (e.g., acne, eczema) | 241 (59.2%) |
| SPF level preference | 204 (50.1%) |
| Cosmetic feel (e.g., non-greasy, mattifying, fragrance-free, tinted) | 235 (57.7%) |
| **Which sunscreen formulation do you commonly recommend for clients**<br>**with oily or acne-prone skin?** |  |
| Spray | 50 (12.3%) |
| Gel | 225 (55.3%) |
| Stick | 14 (3.4%) |
| Lotion | 77 (18.9%) |
| Foam | 14 (3.4%) |
| I'm not sure | 27 (6.6%) |
| **Which sunscreen formulation do you commonly recommend for clients with dry skin?** |  |
| Spray | 23 (5.7%) |
| Gel | 26 (6.4%) |
| Stick | 13 (3.2%) |
| Lotion | 54 (13.3%) |
| Cream | 271 (66.6%) |
| I'm not sure | 20 (4.9) |
| **Which sunscreen formulation do you commonly recommend for clients with sensitive skin?** |  |
| Spray | 81 (19.9%) |
| Gel | 58 (14.3%) |
| Stick | 17 (4.2%) |
| Lotion | 105 (25.8%) |
| Cream | 77 (18.9) |
| I'm not sure | 69 (17.0%) |
| **Do you advise parents to apply sunscreen for their children?** |  |
| Yes | 339 (83.3%) |
| No | 68 (16.7%) |
| **Do you provide patients with guidance on sunscreen application?**<br>**(e.g., reapplication after swimming, amount to apply, etc.)** |  |
| Yes | 373 (91.6%) |
| No | 34 (8.4%) |
| **Would you be interested in additional training or resources to**<br>**improve your ability to guide patients on sunscreen formulations?** |  |
| Yes | 378 (92.9%) |
| No | 29 (7.1%) |

Attitude and practice scores (p<0.001) were strongly impacted by age, with the 30–39 age group exhibiting the greatest attitude. Attitude and practice were influenced by gender (p<0.001), with women scoring higher than men. Also, both (p=0.007) were significantly influenced by years of experience; pharmacists with 11–15 years had the greatest attitude ratings. Moreover, attitude and practice (p=0.003) were influenced by marital status, with married participants displaying better. Furthermore, the score was significantly higher (p<0.001) among graduates from public colleges.

### Predictors of better knowledge, attitude and practice towards usage of sunscreen

Participants with an experience 5-10 years were less likely to have better knowledge of sunscreen compared to others (p<0.05). On the other side, participants who work in independent community pharmacy showed higher odds of being knowledgeable of sunscreen compared to others (p<0.05).

Participants aged 30-39 years, those who have BSc in Doctor of Pharmacy, those with years of experience above 5 years, those who are married and those who graduated from public university showed lower odds of being knowledgeable of sunscreen types and Fitzpatrick scale compared to others (p<0.05).

Participants aged 30-39 years, those with postgraduate studies, those with 11-15 years of experience, those who work in academia, those who are married, and those who graduated from public university were more likely to show positive attitude towards the use of sunscreen (p<0.05).

Table 5 presents the odds ratios and (95%) confidence intervals for factors associated with knowledge, knowledge of sunscreen types and Fitzpatrick scale and attitude towards use of sunscreen.

**Table 5.**
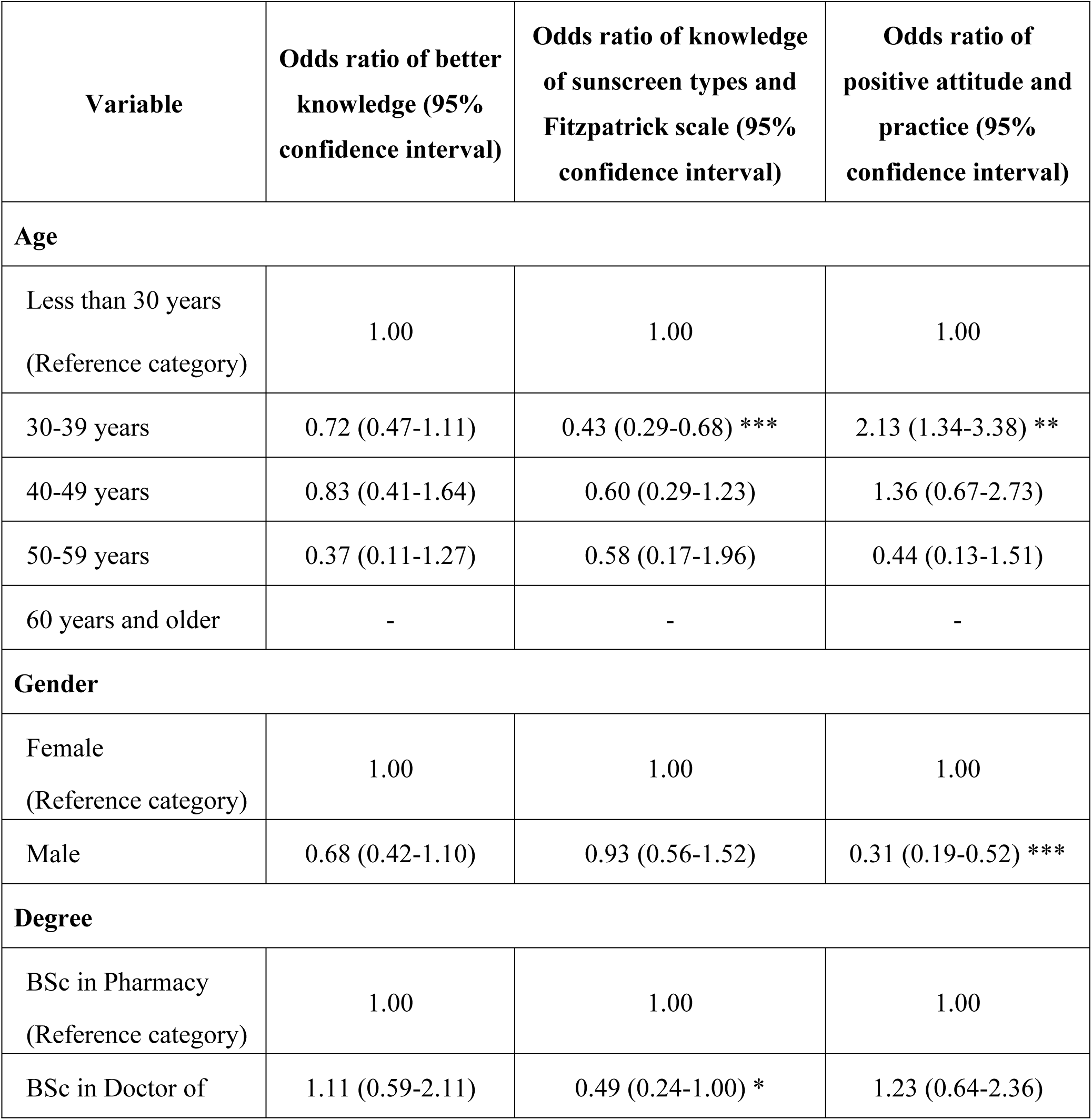
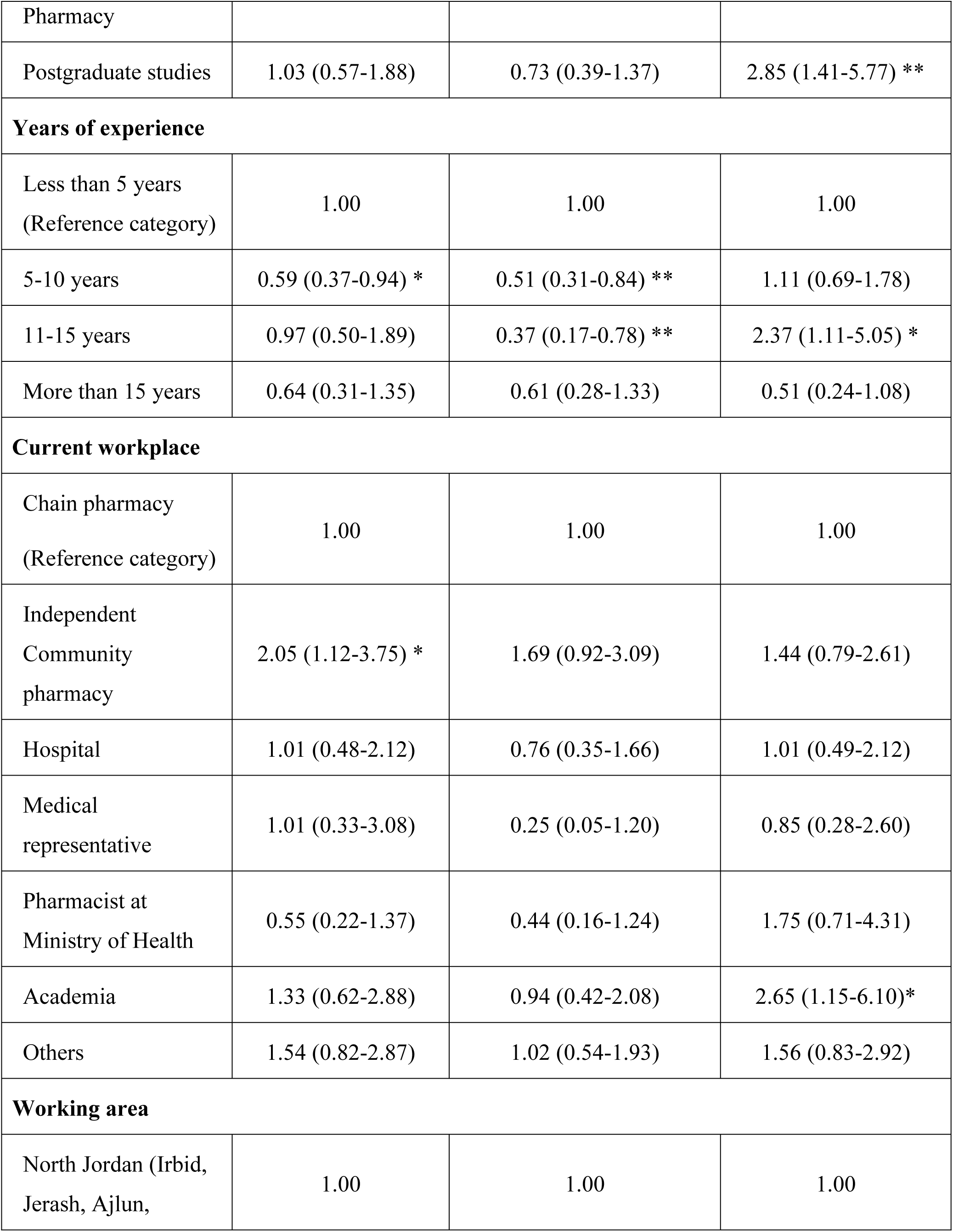
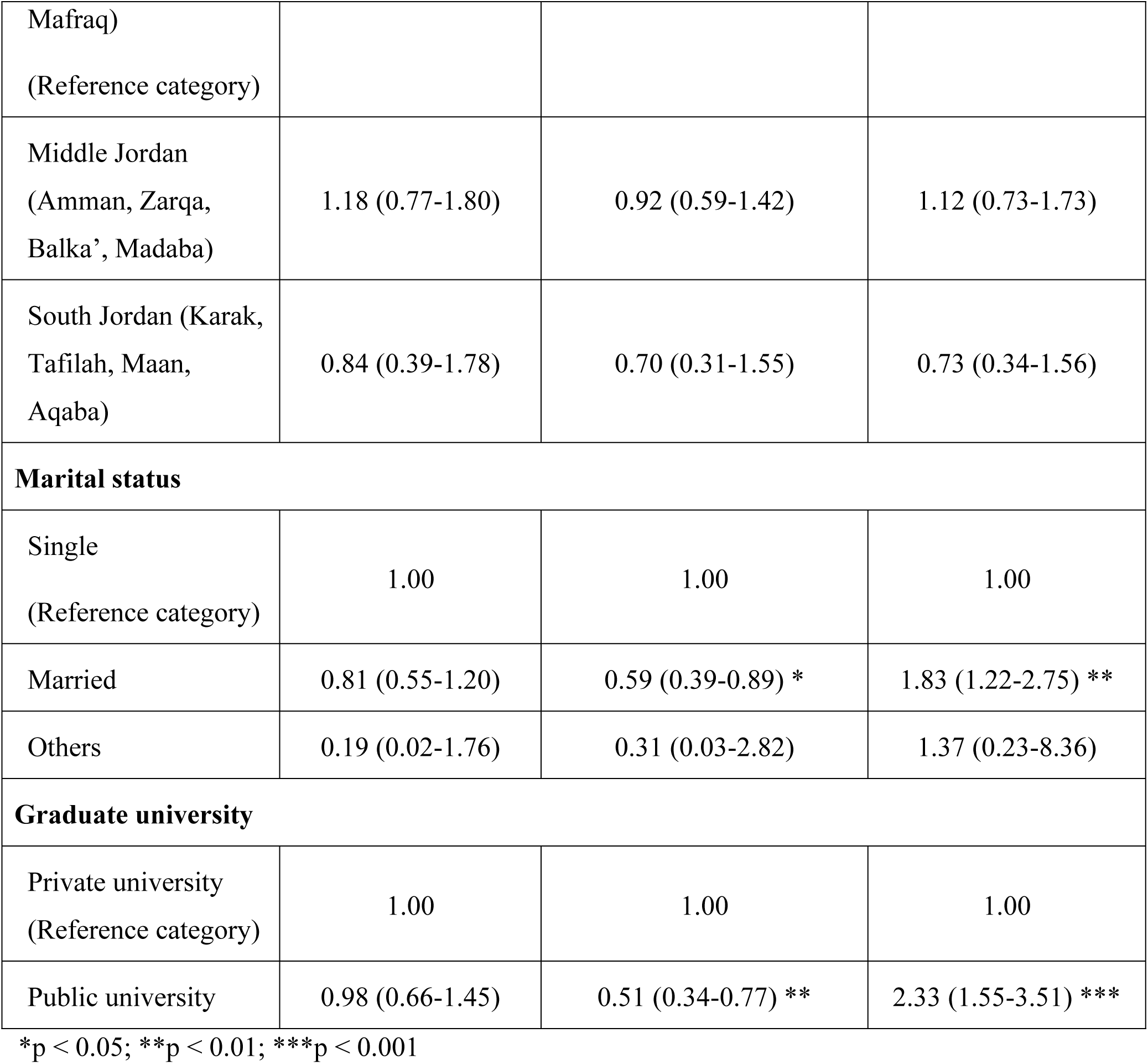
Predictors of better knowledge, knowledge of sunscreen types and Fitzpatrick scale and attitude and practice towards use of sunscreen.

## Discussion

To our best knowledge, this study is the first to assess knowledge, attitude, practices, and perceptions regarding the use of sunscreen among pharmacists in Jordan. While previous studies have evaluated the Knowledge and practices regarding sunscreen use among university students [11], and among Dermatology Patients at University Hospital. [12]

According to our study, the knowledge score between the participants shows a statically significant level (p=0.011), which reflect a high awareness of sunscreens between pharmacists. This goes aligned with the study that assess the knowledge of pharmacist on sunscreen and skin cancer. [13] Our study presented that pharmacists are familiar with the term SPF (70.3%) followed by Somewhat familiar (25.3%), Furthermore, in this study the pharmacists who determine how SPF long it would take for your skin to burn with sunscreen compared to without the use was around (75.9%) also community pharmacists in Jeddah, Saudi Arabia showed that (83%) are knower that the SPF is a Sun protection factor. [14] However, most of the pharmacists in Mashad have inadequate knowledge about the sunscreens and moisturizers and their formulations. [15] According to these results, it indicates that pharmacists have a good understanding of the term SPF. This is a good indicator of the pharmacists’ level of knowledge. Therefore, it is important to adhere to the appropriate level of education to increase awareness and knowledge of these terms. Moreover, in our study the results presented that pharmacists who answered that SPF measure the level of protection against UVA rays are around (44.5%), while who answered that SPF measure the level of protection against UVB rays are around (47.7%). Compared to community pharmacists in Jeddah, Saudi Arabia (72.0%) who answered that SPF measure the level of protection against UVB rays followed by (19.0%) who answered that SPF measure the level of protection against UVB, and UVA, and UVC. [14] These results indicate that pharmacists in Jordan need to increase their knowledge and awareness of the differences between sun rays and how sunscreen works. This can be implemented by giving more lectures on cosmetics at universities and providing specialized courses in this field.

Furthermore, our study the results presented that the percentage of pharmacists who answered that SPF level provides (97%) protection is 50 was around (54.1%) while the percentage of pharmacists in the United Arab Emirates (UAE) who answered that the optimal SPF for use is 15 −20 was (100%). [16] Compared to pharmacists in Jeddah, the results showed that broad-spectrum sunscreens with SPF less than 15 are not used to prevent sunburn only. [14] These different results indicate a lack of knowledge about the sunscreen subject and more focus should be given on this subject by giving lectures related to cosmetic matters and sunscreens Although there is no work that directly evaluate pharmacists’ familiarity with the Fitzpatrick scale, our study is the first study that highlighted the knowledge of sunscreen types and the ingredients of each type of according the Fitzpatrick scale of skin types among pharmacist, the knowledge obtains a significant score (p=0.011), and about (63.0%) of pharmacists responded that they either always or often explain the differences between sunscreen formulations to their clients. Additionally, (85.0%) recommend the sunscreen dosage form according to the skin’s type, (60.7%) according to lifestyle or activity level (e.g., sports, beach, everyday use), (59.2%) according to skin concerns (e.g., acne, eczema) and (57.7%) according to cosmetic feel (e.g., non-greasy, mattifying, fragrance-free, tinted).

A study illustrates the importance of knowing the individual skin phototypes among healthcare providers plays a vital role in tailoring sunscreen recommendations and prevention strategies. [17] Moreover the need to be acknowledged about how sunscreen agents are working for UV ration protection and how to know the type of skin which assess in understanding how each type of the 6 types of Fitzpatrick scale reacts once exposed to sunlight. [18]

Regarding prescribing dosage from sunscreen according to the skin type; (55.3%) and (66.6%) of pharmacists responded correctly that gel form and cream form are the recommended forms for clients of oily skin and dry skin respectively, while (25.8%) and (19.9%) recommend lotion form and spray form, respectively, for sensitive skin. These results show that pharmacists should be educated more about the correct dosage form to be prescribed according to skin type either throughout university education or by further training courses conducted by companies producing sunscreen agents as part of pharmacists’ continuous professional development, and (83.3%) of pharmacists advise parents to apply sunscreen for their children.

The study results showed that (86.0%) of pharmacists apply sunscreens for themselves and (94.3%) recommend usage of sunscreen to others. (81.6%) of pharmacists recommend users to apply chemical sunscreens 15-30 minutes before sun exposure every 2-4 hours (72.2%). These results agree with Kurban, N. A. et.al who studied the awareness of cosmeceutical including sunscreen agents among pharmacists in Saudi Arabia who found that (90.0%) of pharmacists think that sunscreens should be applied 15-20 minutes before sunlight exposure and (92.0%) think that sunscreens should be reapplied every 2 hours. [14]

In our study the vast majority (94.6%) responded that sunscreens are effective in sunburn prevention and (72.2%) of pharmacists believe that sunscreens are effective in preventing skin damage, this result is like a study conducted in Malaysia showed that are pharmacists aware that sunscreen is effective in preventing sunburn, skin aging. [19] The additional advantages of sunscreen usage comprise prevention of sunburn, ageing, pigmentary disorder and skin cancer. [20]

Intriguing, (43.5%) of pharmacists think that sunscreens can lead to Vitamin D deficiency, it was reported that sunscreens could affect vitamin D photoproduction in case its applied at the tested concentration. 1/SPF of UV radiation is transmitted into skin if applied at concentration of 2mg/cm2 (the tested level of protection) while users commonly apply 0.5 mg/cm2 which is unlikely to cause vitamin D deficiency. Also applying sunscreen agents extend outdoor spending period that increase exposing to UV radiation. [21]

Most pharmacists (89.7%) considered sunscreen are safe to use during pregnancy which align with Lim, et al. who reviewed the benefits of sunscreens usage during pregnancy as diminishing pigmentation changes from UV exposure during pregnancy. [22] (79.1%) believe that sunscreens are safe for babies older than 6 months, this aligns with the recommendation of U.S. Food and Drug Administration under children’s health topic that sunscreens should not apply for babies less than 6 months year old. [23]

Most pharmacists (91.6%) replied that they provide patients with guidance on sunscreen application as reapplication after swimming and amount to apply. Moreover (92.9%) of pharmacists are interested in additional training or resources to improve your ability to guide patients on sunscreen formulations.

According to our study, participants who work in independent community pharmacy showed higher odds of being knowledgeable of sunscreen compared to others (p<0.05). This is in line with a study [19], which showed that pharmacists described a higher rate of right answer according to knowledge of sunscreen than physicians even the statistical significance not reached (p=0.008).

Gender was also a factor that influenced the towards sunscreens, female participant in our study scoring higher than male (p<0.001), reflecting their greater use of sunscreen. This result goes along side with other study conducted in southern Sweden showed that the female gender was used the sunscreen extensively in comparison to men. Also, with increase the age the level of protection increased. (p<0.001). [24]

Interestingly, participants aged 30-39 years, pharmacists holding Doctor of Pharmacy degree, those with years of experience above 5 years, married and those who graduated from public university showed lower odds of being knowledgeable of skin and sunscreen types compared to others (p<0.05). similarly to a prior study illustrated that the Socio-demographic have correlation with the use of sunscreens are gender (female more than male), higher status, higher levels of education, skin type IV and married individuals. [25] These results highlighted the influence of demographic and professional characteristics on knowledge, attenuate and practice, and further indicated the necessitates the need for health education program.

Importantly, [26] pointed out that the curricula of the American College of Clinical Pharmacy, and the American Society of Consultant Pharmacists discovered negligible coursework on dermatology-related diseases and treatment. Showing that most pharmacists are not fully trained in counseling patients with dermatological conditions. This lack of specialized education courses in this field at universities, that can explain the limited knowledge of skin types and sunscreen formulations among pharmacists. Therefore, incorporating structured dermatology-focused content into pharmacy education could bridge this gap and improve pharmacists’ competence in sun protection counselling.

### Conclusion

This study concluded that a significant percentage of pharmacists are informed about sunscreen agents and their knowledge about sunscreen was lower than their attuite and practice. This lead us that pharmacist needs more specialized courses during practice for their continuing professional development to improve the knowledge and enhance the practice more. Alongside with that, pharmacy students as well need more courses that highlight comprehensive information about sunscreen agents in their courses.

## Data Availability

All relevant data are within the manuscript and its Supporting Information files.

## Acknowledgment

We would like to acknowledge Princess Nourah bint Abdulrahman University Researchers Supporting Project, Princess Nourah bint Abdulrahman University, Riyadh, Saudi Arabia.

## Notes

### Competing Interest Statement

The authors have declared no competing interest.

### Funding Statement

Yes

### Author Declarations

The approval from the Deanship of Scientific Research and Graduate Studies at Jerash University, with the ethical approval number (2025/2024/2/7)

